# The trials of interpreting clinical trials - A Bayesian perspective Colchicine in secondary cardiovascular prevention

**DOI:** 10.1101/2025.05.04.25326946

**Authors:** James M Brophy

**Affiliations:** McGill University Health Center Centre for Health Outcomes Research (CORE) 5252 Boul. de Maisonneuve West, Montréal QC H4A 3S5; Department of Medicine, McGill University, Montréal, QC; Department of Epidemiology, Biostatistics and Occupational Health, McGill University, Montréal, QC

## Abstract

**Objectives:** Evidence based medicine (EBM) places systematic reviews and meta-analyses, at the top of the evidential pyramid. Bayesian methods may assist in better understanding uncertainties and improve interpretations and harmonization.

**Design:** A 2022 meta-analysis concluded that colchicine reduced the cardiac risk in secondary prevention. Nevertheless, a large, RCT (CLEAR) continued to randomize acute patients to colchicine or placebo and in 2025 published their findings of no benefit. Bayesian sequential analyses and hierarchical meta-analysis can inform the decision to complete this trial and augment the nuances surrounding its interpretation.

**Setting:** RCTs of coronary artery disease (CAD) patients with an acute coronary syndrome admission undergoing percutaneous coronary intervention (PCI).

**Interventions:** Randomization to colchicine or placebo.

**Main outcomes:** The primary outcome was major adverse cardiovascular events (MACE), a composite of death from cardiovascular causes, recurrent myocardial infarction, stroke, or unplanned ischemia-driven coronary revascularization.

**Results:** A published 2022 meta-analysis suggested a statistical MACE decrease with colchicine (RR 0.73 [95% confidence interval (CI) 0.62, 0.86]), but a Bayesian reanalysis showed a 95% credible interval (95% CrI 0.26, 1.70) for the next study, justifying continuing the CLEAR tiral. CLEAR results were eventually interpreted as “negative” (HR, 0.99; 95% confidence interval [CI], 0.85 to 1.16). Bayesian sequential re-analyses using a vague prior (i..e. result dominated by CLEAR), an all-inclusive prior (based on the previous meta-analysis), and a focused prior (considering only the largest and most similar previous RCT) showed 58%, 100% and 92% probabilities respectively of a MACE decrease with colchicine. The probabilities of clinically meaningful decreased, based on > absolute 15% MACE reduction, were more modest, between 2% - 41%.

**Conclusions:** Bayesian analyses offer advantages in clinical trial design and interpretation. The worked example strongly suggests some benefit for colchicine in secondary cardiovascular prevention, but it is unlikely to be of clinical importance.

## Introduction

The evidence based medicine (EBM) paradigm places systematic reviews and meta-analyses, ideally of randomized clinical trials, at the top of the evidential pyramid(1). While symbolically straight forward, there are numerous intricacies in properly assessing the quality and interpretation of individual RCTs and meta-analyses. Reporting guidelines[(2)](3) have been proposed to hopefully improve quality. However, there remain difficulties in scientific communication leading to misinterpretations of both clinical trials(4) and meta-analyses(5). The underlying causes are a multifactorial combination of a failure to systematically integrate previous and current trials[(6)](7), an under appreciation and estimation of uncertainties(8) and cognitive biases associated with the standard null hypothesis significance testing paradigm(9).

The incremental advantages of Bayesian approaches over more traditional methods to decide when additional research is required, how to interpret the new results, how to synthesize and communicate the totality of the evidence will be presented. At each stage the incremental benefits of the Bayesian approach over the traditional statistical approach will be highlighted. While additional statistical insights are demonstrated, these remain ancillary to the crucial element of clinical judgement. Analogously to statistical significance not equating unreservedly to causality, clinical judgement must precede statistical analyses in deciding what is a “reasonable” research question, design, analysis and when are different studies “combinable”.

The importance of these issues will be demonstrated with the contemporary example of colchicine in the prevention of cardiovascular events in patients with symptomatic (acute) coronary artery disease (CAD) who have had acute percutaneous coronary interventions (PCI) with particular attention to the execution and interpretation of the most recent study(10).

## Methods

We initially reproduced the 2022 systematic review and meta-analysis(11) of colchicine for patients with symptomatic CAD who underwent acute PCI, published before the completion of the 2025 CLEAR(10) trial. This not only assured reproducibility of prior evidence but also permitted the addition of the valuable[(12)](13), but missing, prediction interval(11). The prediction interval are derived as follows

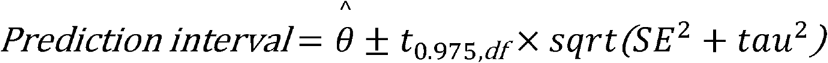

where:

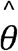 is the pooled log(RR),

SE is its standard error,

*τ*^2^ is the between-study variance, and

*t*_0.975,*df*_ is the critical value from the t-distribution with k−2 degrees of freedom.

By explicitly quantifying the uncertainty in the mean with the estimated between trial heterogeneity (*τ*^2^) the prediction interval shows the plausible range for a new study’s true effect rather than just simply reporting the precision of the average pooled effect. Prediction intervals can be back-transformed to the Risk Ratio (RR) scale.

However, frequentist prediction interval calculations assumes the parameters are perfectly known and fixed, thereby limiting their natural variability. The limitations and under-estimation of frequentist random effects meta-analytical models has previously been noted(14). Bayesian hierarchical random-effects model of aggregated study-level data acknowledge the uncertainty with each observed study effect estimate, *y*_*i*_, modeled as 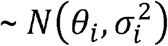, where *σ*_*i*_ is the known standard error for each study.

The latent true *study*_*i*_ effect, denoted by *θ*_*i*_, is also assumed to vary across studies and is itself modeled as ∼ N(*µ, τ*^2^), where *µ* is the overall average effect, and *τ* is the between-study standard deviation, also known as the heterogeneity. Vague priors can be assigned to these parameters; a standard normal prior for *µ* ∼ N(0,1), and a halfnormal prior (via *τ* ∼ normal(0, 1) with *τ* >= 0) for *τ*. Together, these define a normal-normal hierarchical model, where partial pooling occurs such that individual study estimates are shrunk toward the overall mean, with the degree of shrinkage determined by *τ*. This model supports posterior inference on µ, study-specific effects *θ*, and provides a prediction interval for a future study by simulating from the posterior predictive distribution, while acknowledging the uncertainty of the model parameters. Typically Bayesian prediction intervals by more realistically accounting for parameter uncertainty will therefore be wider than their frequentist counterparts. In addition to reproducing the original 2022 meta-analysis(11), we also updated it in a Bayesian framework using the same original search strategy but updated to April 20 2025.

Rather than concentrating on the prediction interval for the next study, interest may center on the interpretation of the most recent study(10), but not as an isolated entity as done in the original publication, but rather in the context of what was previously known. This sequential updating of prior evidence with new data is the essence of Bayesian inference and mirrors human learning. The binary observed summary statistic *y*, the log risk ratio from CLEAR(10) can be represented by a binomial distribution. but given the large sample size, the the log risk ratio is equally well approximated by a normal distribution, modeled as *y* ∼ N(*µ, σ*^2^), with assumed known standard error *σ*. A normal prior is again placed on 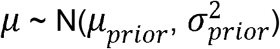. The model returns a posterior distribution for *µ*, combining prior information and observed data in accordance with Bayes’ theorem. Identical results are obtained with a binomial instead of a normal distribution of the data (results not shown).

As priors may be considered subjective, multiple priors or a community of priors, can assess concerns of subjectivity and test the robustness of the conclusions. Adopting the CLEAR investigators’ published viewpoint of trial interpretation independent of prior knowledge implies a vague prior so that the posterior probability distribution is dominated by the observed CLEAR data. However, CLEAR(10) investigators also acknowledged previous evidence, particularly the COLCOT(15) trial. It therefore seems appropriate to consider two additional priors, COLCOT(15) and one that uses all prior relevant evidence as summarized in the previous meta-analysis(11). These three priors can be specified as follows;

‐ a vague prior (N(0, 2)) for the mean summary statistic, log(RR), which allows CLEAR data to dominate and approximate the published frequentist result
‐ a fully informative mean summary prior using all previous data as summarized by the prediction interval of the earlier meta-analysis(11)
‐ a “focused” mean summary prior based on the largest, highest-quality, most similar previous RCT, COLCOT(15), which also has the most positive results

All analyses were preformed within the R ecosystem(16) and the RStudio Integrated Development Environment(17). The reproduction of the frequentist random effects was performed with the metafor package(18). The Bayesian hierarchical meta-analysis to synthesize the evidence estimated by the pooled log risk ratio used the Stan programing language(19) with sampling of the posterior distribution via the cmdstanr package(20). Although the sequential updating approach defines a conjugate model — meaning the posterior is also normally distributed and could be derived analytically — we have not used this closed-form solution. Instead, we again used numerical methods(19) specifically, Hamiltonian Monte Carlo sampling of the posterior distribution of *µ*, allowing maximum flexibility and extendibility (e.g., to non-conjugate or hierarchical models). All statistical code is available at https://github.com/brophyj/colchicine

## Results

### Previous studies

The 2022, pre-CLEAR(10), systematic review and meta-analysis(11) of the effects of colchicine in patients with symptomatic acute CAD who underwent PCI reported reduced MACE outcomes (RR 0.73 (95% CI 0.61 to 0.87); p=0.0003) with minimal heterogeneity (*I*_2_=6%). Our calculated frequentist pooled summary estimate was identical to the published result and our frequentist calculated prediction interval was only marginally widened (0.60 - 0.90) due to the limited observed between study heterogeneity and the falsely high precision associated the commonly employed DerSimonian–Laird estimator(14). Based on the upper 95% CI of this model, the next colchicine study was predicted to almost certainly have at least a 10% reduction in cardiovascular outcomes. However, the fragility of that conclusion becomes apparent if a different model for the between study variation with heavier tails than the current *t(*_*n-k*_)_*df*_ choice is assumed. For example with a critical value of *t*_0.975,2_ the upper 95% CI for the predicted interval becomes 1.04, suggesting the need for further studies to reduce the uncertainty of the summary estimate.

Moreover with only a few studies, treating *τ* and *µ* as known and fixed quantities can seriously underestimate uncertainty. A Bayesian model averages over the joint posterior distribution of these two parameters better reflecting the uncertainty in how effects vary between studies (heterogeneity), while also accounting for uncertainty in the pooled average effect itself. This results in a wider and more realistic prediction intervals (0.35 - 1.29) that better captures the expected range of effects for a new study (Table 1). Unlike the frequentist analysis, this approach confirms the need for additional research to better define any beneficial colchicine effect.

**Table 1.**
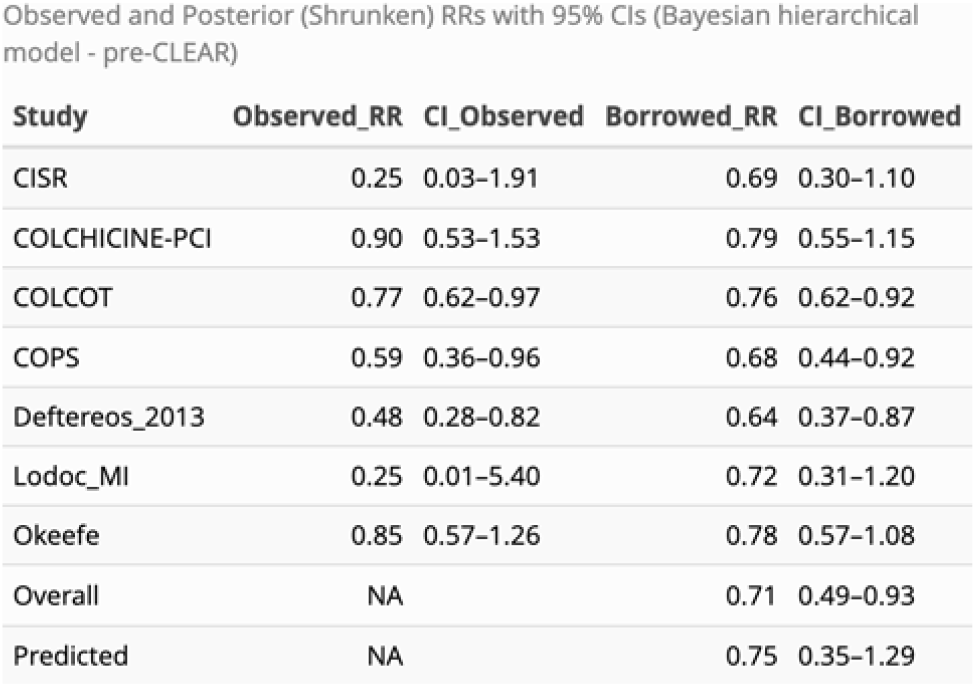

### The new evidence

The updated 2022 meta-analysis(11) search strategy to April 20 2025 (see PRISMA flowchart (to April 2025) in the supplemental material) identified only one additional RCT, the CLEAR(10) trials which randomized post myocardial infarction patients patients between February 1, 2018, and November 8, 2022 to colchicine (n = 3,528) or placebo (n = 3,534) immediately following PCI. The primary outcome was MACE, a composite of cardiovascular death, MI, stroke, or ischemia-driven revascularization. With a median 3 year follow-up, there were 9.1% events in the colchicine group and 9.3% in the placebo arm (HR 0.99, 95% CI 0.85-1.16). Given the high quality study design, execution, and large number of primary outcome events (n=649), the authors reasoned that the probability of a spurious result was low. Thus, they concluded “… treatment with colchicine, when started soon after myocardial infarction and continued for a median of 3 years, did not reduce the incidence of the composite primary outcome.”(10)

CLEAR authors acknowledge that the most comparable previous study was COLCOT(15), a randomized trial of 4745 patients within 30 days of an acute myocardial infarction, who received the same interventions with essentially the same composite primary enpoint and a median 22.6 months follow-up. COLCOT(15) had fewer total primary-outcome events (n=301), but nevertheless found a 23% relative risk reduction (HR 0.77, 95% CI 0.61 - 0.96). In the CLEAR discussion(10), there was no attempt to explain these “differences” other than observing that two colchicine trials in stroke patients[(21)](22) also showed no benefit with colchicine and that CLEAR was a larger trial, presumably with an improved precision of any treatment effect.

Should clinicians adopt the CLEAR(10) investigators’ view that the larger trial is to be believed and that the previous evidence from the 4745 COLCOT(15) subjects or the 6660 meta-analysis(11) subjects should simply be forgotten? The CLEAR PI was more explicit in an interview following an oral presentation of their findings stating “I was a believer in colchicine but after CLEAR I decided to stop it in my parent”(23). Presumably his prior belief wasn’t universally shared, or at least believed at a clinically significant threshold, as otherwise equipoise would have been lacking to proceed with the CLEAR trial. In any case, this dichotomization of thinking for accepting or rejecting null hypotheses significance testing and for clinical decision making, while common among clinicians, is known to favour misinterpretations(9).

### Sequential evidentary updating - A Bayesian approach

The above approach where decisions are conditioned on the comparison of p values (probability of the observed or more extreme data | null hypothesis) to prespecified type I errors, typically 0.05 is standard in medical research. Unfortunately this approach may result in the aforementioned cognitive errors[(24)](25)(26). On the other hand, a Bayesian approach provides the information clinicians are actually seeking, namely the probability that the hypothesis is true given the observed data, the so called “inverse probability”. This posterior distribution, derived from a weighted combination of a prior belief and the current data, incorporates prior knowledge according to the rules of probability via Bayes Theorem, and minimizes interpretative errors(6). A further advantage of probability distributions is they are not restricted to specific point estimates but permit calculations for multiple different cutpoints or intervals. For example, one might be particularly interested in probabilities exceeding a clinically meaningful thresholds for benefit or harm. Such clinical cutpoints can be individually selected but for demonstration purposes with the colchicine example a benefit threshold of RR < 0.9 and harm threshold of RR > 1.1 has been chosen.

Figure 1 graphically displays the three priors and the three updated posteriors after the incorporation of the CLEAR data. The informative posteriors are a weighted average of the individual priors and CLEAR data (blue distribution) with weights proportional to their precisions and therefore a shifting of the priors towards the CLEAR data. Informative posterior distributions also show the expected improved precision with a narrowing distributions due to the inclusion of the new data. The posterior distribution with a vague prior corresponds to the CLEAR data alone, the so-called likelihood.

**Figure 1.**
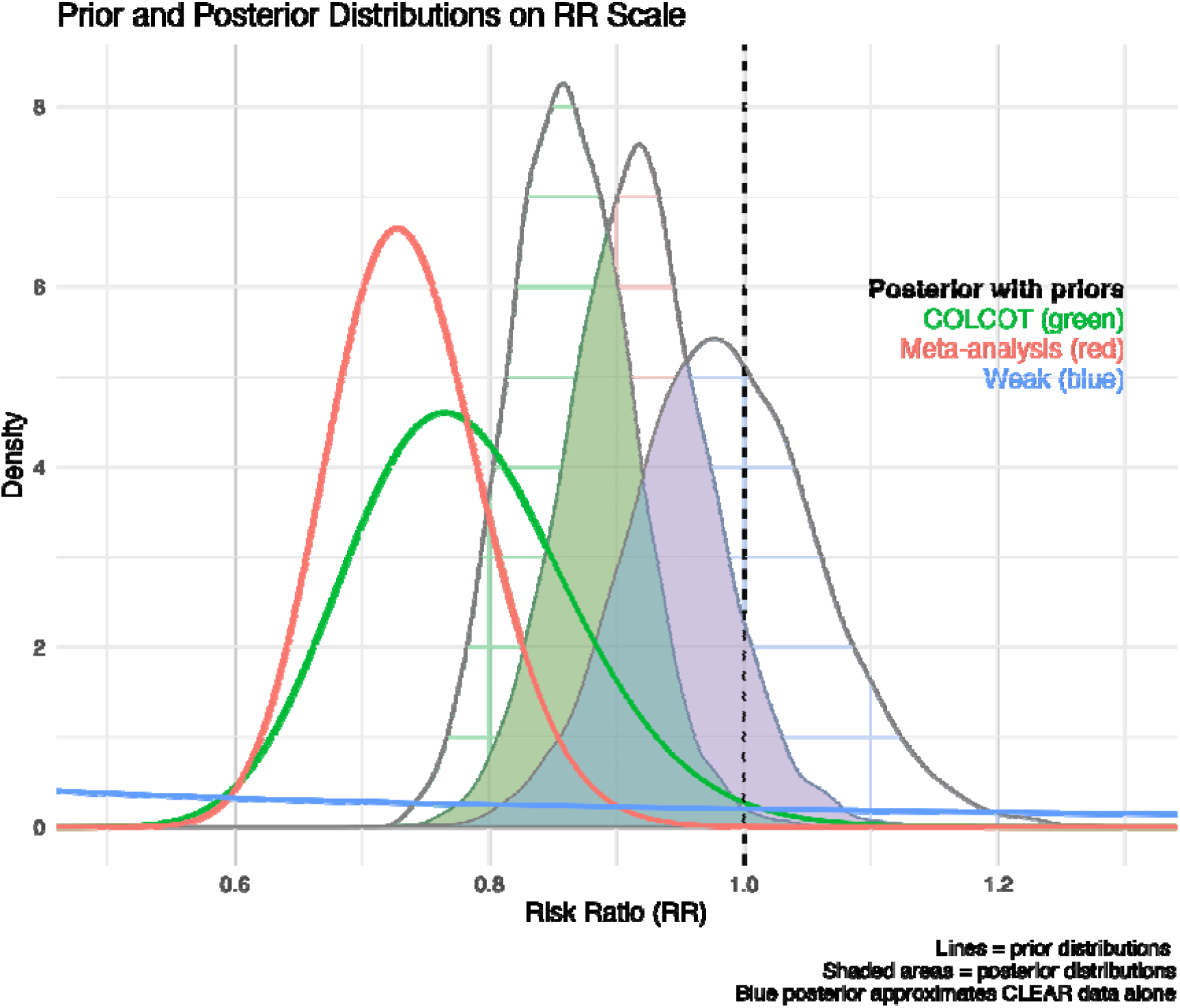

Numerical results are derived from integration of the area under the posterior curves (AUC) to the left of any selected threshold and are given in Table 2. The posterior results using informative priors suggests a 92% to 100% probability of decreased MACE outcomes (AUC < 1). Moreover, these analyses suggest a possible 12% to 41% probability that the MACE decrease is at least 15% (AUC < 0.85). Regardless of the chosen prior, there is virtually no chance that colchicine increases cardiac risk by a clinically meaningful amount (AUC < 1.15).

**Table 2.** Posterior Risk Ratio (RR) summaries and probabilities (P) for CLEAR trial under different priors

| prior | Mean [95% CrI] | P(RR < 0.8) | P(RR < 0.85) | P(RR < 0.9) | P(RR < 1.0) | P(RR < 1.15) |
| --- | --- | --- | --- | --- | --- | --- |
| COLCOT prior | 0.92 [0.81, 1.04] | 0.02 | 0.12 | 0.39 | 0.92 | 1.00 |
| Meta-analysis prior | 0.86 [0.77, 0.96] | 0.09 | 0.41 | 0.79 | 1.00 | 1.00 |
| Weak prior | 0.99 [0.85, 1.14] | 0.00 | 0.02 | 0.11 | 0.57 | 0.98 |

### Bayesian Meta-analysis

Updating prior beliefs is temporally consistent with data availability and mirrors human sequential learning. A different perspective that also combined all available data but concentrates on the summary result and predictability of the next future study is provided by meta-analytical techniques. This approach ignores data temporality and is problematic when the total number of trials is small. A Bayesian random effects (hierarchical) meta-analysis is preferred over the frequentist model, since as discussed above it models all parameter uncertainty. Individual studies are considered part of a larger population of studies and individual study estimates are “pulled” and “shrunken” toward the overall mean effect. Hierarchical models represent a compromise between complete pooling (fixed effect) or no pooling (complete study independence). Hierarchical models thereby account for both within and between study variability producing pooled mean estimates that integrate information from different studies while acknowledging their variability. These models also can provide the predictive interval for the next study from the super population of possible studies.

The Forest plot for the updated Bayesian meta-analysis with CLEAR(10) in shown in Figure 2 with numerical results in Table 3. The figure demonstrates the shrinkage of each observed trial result towards the global mean. It also shows a point estimate for global mean that is close to the pre-CLEAR pooled value but with wider 95% CI and the prediction intervals due to the increased between trial heterogeneity

**Table 3.** Observed and Posterior (Shrunken) RRs with 95% CrIs (Bayesian hierarchical model - post-CLEAR)

| Study | Observed_RR | CI_Observed | Shrunken_RR | CrI_Shrunken |
| --- | --- | --- | --- | --- |
| CISR | 0.25 | 0.10–0.64 | 0.51 | 0.21–0.87 |
| COLCHICINE-PCI | 0.90 | 0.55–1.49 | 0.84 | 0.55–1.28 |
| COLCOT | 0.77 | 0.65–0.92 | 0.77 | 0.65–0.91 |
| COPS | 0.59 | 0.44–0.79 | 0.61 | 0.46–0.79 |
| Deftereos_2013 | 0.48 | 0.37–0.63 | 0.52 | 0.39–0.66 |
| Lodoc_MI | 0.25 | 0.02–3.63 | 0.69 | 0.25–1.38 |
| Okeefe | 0.85 | 0.60–1.20 | 0.82 | 0.59–1.11 |
| CLEAR | 0.99 | 0.85–1.16 | 0.97 | 0.83–1.13 |
| Overall | NA |  | 0.69 | 0.47–0.94 |
| Predicted | NA |  | 0.78 | 0.26–1.70 |

**Figure 2.**
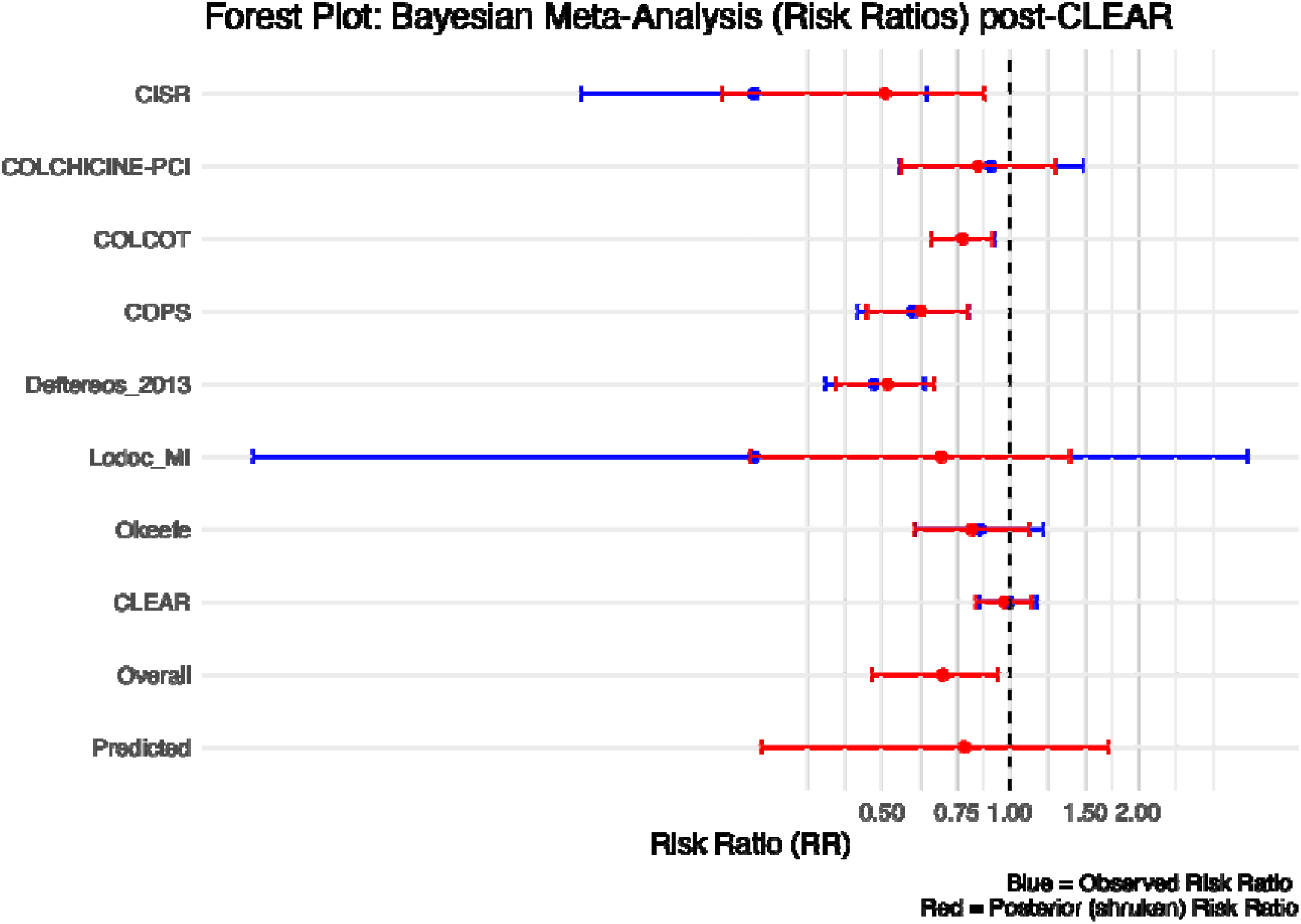

## Conclusion

While the published conclusion of the CLEAR(10) trial was that colchicine did not reduce the incidence of MACE, this Bayesian re-analysis with two well justified informative priors suggests there is a substantial probability (57% to 100%) that colchicine does reduce MACE, although the probability that this benefit exceeds a clinically meaningful 10% reduction is more modest (between 39% to 79%). More meaningful risk reductions are less likely but not completely impossible (2% to 41% probability of a 15% reduction). This Bayesian approach allows a more complete and nuanced examination not only of the recent CELAR(10) study on on its own merit but also in the context of existing evidence. While it may seem disconcerting that after 13,000 randomized patients we do not have an unequivocal answer to the research question, this nonetheless reflects reality. These results should not be interpreted as a rejection of the colchicine hypothesis as implied by CLEAR(10) but rather alternatively as showing that while a large clinical benefit is unlikely the possibility of a modest colchicine clinical benefit has not been definitively excluded.

This study also highlights the importance of considering prediction intervals and a Bayesian approach in meta-analysis. Had the CLEAR investigators relied on the pooled summary estimate from the 2022 frequentist meta-analysis(11), they would have prematurely and inappropriately stopped their trial for ersatz colchicine efficacy. Prediction intervals are important in meta-analyses to assess the likely range of a future study. However as this example shows, frequentist methods often under-estimate the uncertainty associated with these intervals and may lead to inappropriate over confident conclusions.

Given that CLEAR(10) and COLCOT(15) were both well designed, well executed trials and published in the same esteemed medical journal, its seems absurd to ignore either of them. A fundamental question is are these two study results really different? First and most importantly from a clinical perspective the trials examine the same intervention and outcomes in very similar populations. From a statistical perspective the results aren’t radically different with overlapping 95% CIs. It is only if each trial is assessed with the inappropriate dichotomous statistical significance lens (p < or > 0.05) that the two trials appear superficially different. The key insight being “In making a comparison between two treatments, one should look at the statistical significance of the difference rather than the difference between their significance levels”(27). The Bayesian statistical lens sharpens this perspective.

Clinicians are regularly faced with “conflicting” trial evidence. However if the trials are of equal high quality, often these conflicts are illusory arising from the improper comparisons of statistical significance. Systematic reviews and meta-analyses of prior evidence are now de riguer before a study is considered for peer review funding. Yet there is no similar mandate for evidence synthesis upon trial completion. Indeed current incentives perversely favour each trial being individually interpreted(28). However, as demonstrated herein, this approach can lead to vacillating beliefs that do not respect the laws of probability and consequently may not align with the true state of knowledge. Bayesian techniques can address these issues thereby raising the quality of clinical trial interpretations.

Consider the colchicine “believer” prior to CLEAR but not afterwards. If COLCOT was responsible for the initial positive prior belief, this analysis suggests after CLEAR there remains a 92% probability of decreased risk with colchicine, a 39% probability that the benefit exceeds a 10% reduction. Of course, if a clinician has an clinical cutpoint for efficacy of RR < 0.80 then it does indeed seem reasonable, based on the totality of the evidence to be a “non-believer” after CLEAR as the probability of a decrease in cardiovascular outcomes of this magnitude is less than 0-2%. However consistency would imply that their prior belief should also be referenced to a probability of RR < 0.80 which for COLCOT was only 60% which seems a modest probability to have become an earlier “believer”. This demonstrates that an intuitive reconciliation of prior and posterior beliefs can be difficult and is not facilitated by dichotomized reasoning. Moreover, clinicians may exhibit an availability bias whereby they are overly influenced by the last trial, particularly if they were intimately involved in it. The probabilistically correct harmonization of all available evidence with a Bayesian analysis can minimize these cognitive errors and an increased emphasis on teaching these methods to clinicians would seem appropriate.

## Data Availability

All data produced in the present study are available

https://github.com/brophyj/colchicine

## Conflict of interest,funding and data sharing statements

There were no funding sources for this work.

The author has no conflicts of interest to report.

The code for these analyses can be found https://github.com/brophyj/colchicine

## Supplemental materila

PRISMA flowchart (to April 2025)

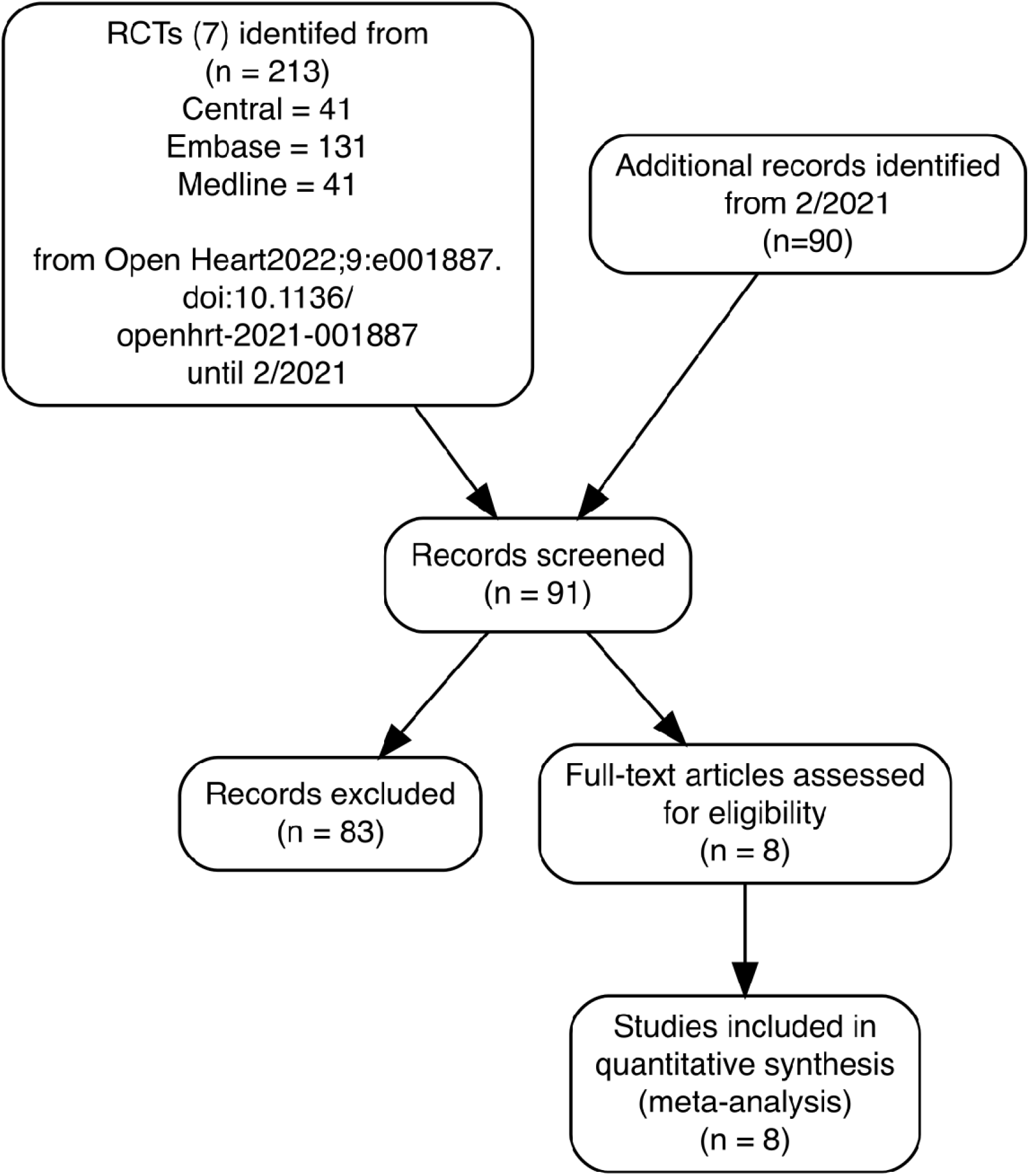

## Notes

### Competing Interest Statement

The authors have declared no competing interest.

### Funding Statement

This study did not receive any funding

### Author Declarations

Only published data was used

## References

1. Guyatt GH, Sackett DL, Cook DJ. Users’ guides to the medical literature. II. How to use an article about therapy or prevention. B. What were the results and will they help me in caring for my patients? Evidence-based medicine working group. JAMA [Internet]. 1994;271(1):59–63. Available from: https://www.ncbi.nlm.nih.gov/pubmed/8258890

2. Moher D, Hopewell S, Schulz KF, Montori V, Gotzsche PC, Devereaux PJ, et al. CONSORT 2010 explanation and elaboration: Updated guidelines for reporting parallel group randomised trials. BMJ [Internet]. 2010;340:c869. Available from: https://www.ncbi.nlm.nih.gov/pubmed/20332511

3. Page MJ, McKenzie JE, Bossuyt PM, Boutron I, Hoffmann TC, Mulrow CD, et al. The PRISMA 2020 statement: An updated guideline for reporting systematic reviews. BMJ [Internet]. 2021;372:n71. Available from: https://www.ncbi.nlm.nih.gov/pubmed/33782057

4. Ioannidis JP. Why most published research findings are false. PLoS Med [Internet]. 2005;2(8):e124. Available from: https://www.ncbi.nlm.nih.gov/pubmed/16060722

5. Pereira TV, Ioannidis JP. Statistically significant meta-analyses of clinical trials have modest credibility and inflated effects. J Clin Epidemiol [Internet]. 2011;64(10):1060–9. Available from: https://www.ncbi.nlm.nih.gov/pubmed/21454050

6. Brophy JM. Bayesian analyses of cardiovascular trials—bringing added value to the table. Canadian Journal of Cardiology. 2021;37(9):1415–27.

7. Spiegelhalter DJ, Myles JP, Jones DR, Abrams KR. Methods in health service research. An introduction to bayesian methods in health technology assessment. BMJ [Internet]. 1999;319(7208):508–12. Available from: https://www.ncbi.nlm.nih.gov/pubmed/10454409

8. Heuts S, Kawczynski MJ, Sayed A, Urbut SM, Albuquerque AM, Mandrola JM, et al. Bayesian analytical methods in cardiovascular clinical trials: Why, when, and how. Can J Cardiol [Internet]. 2025;41(1):30–44. Available from: https://www.ncbi.nlm.nih.gov/pubmed/39521054

9. Rafi Z, Greenland S. Semantic and cognitive tools to aid statistical science: Replace confidence and significance by compatibility and surprise. BMC Med Res Methodol [Internet]. 2020;20(1):244. Available from: https://www.ncbi.nlm.nih.gov/pubmed/32998683

10. Jolly SS, d’Entremont MA, Lee SF, Mian R, Tyrwhitt J, Kedev S, et al. Colchicine in acute myocardial infarction. New England Journal of Medicine [Internet]. 2025;392(7):633–42. Available from: https://www.ncbi.nlm.nih.gov/pubmed/39555823

11. Aw KL, Koh A, Lee HL, Kudzinskas A, De Palma R. Colchicine for symptomatic coronary artery disease after percutaneous coronary intervention. Open Heart [Internet]. 2022;9(1). Available from: https://www.ncbi.nlm.nih.gov/pubmed/34992158

12. Riley RD, Higgins JP, Deeks JJ. Interpretation of random effects meta-analyses. BMJ [Internet]. 2011;342:d549. Available from: https://www.ncbi.nlm.nih.gov/pubmed/21310794

13. IntHout J, Ioannidis JP, Rovers MM, Goeman JJ. Plea for routinely presenting prediction intervals in meta-analysis. BMJ Open [Internet]. 2016;6(7):e010247. Available from: https://www.ncbi.nlm.nih.gov/pubmed/27406637

14. Cornell JE, Mulrow CD, Localio R, Stack CB, Meibohm AR, Guallar E, et al. Random-effects meta-analysis of inconsistent effects: A time for change. Ann Intern Med [Internet]. 2014;160(4):267–70. Available from: https://www.ncbi.nlm.nih.gov/pubmed/24727843

15. Tardif JC, Kouz S, Waters DD, Bertrand OF, Diaz R, Maggioni AP, et al. Efficacy and safety of low-dose colchicine after myocardial infarction. N Engl J Med. 2019;381(26):2497–505.

16. R Core Team. R: A language and environment for statistical computing [Internet]. Vienna, Austria: R Foundation for Statistical Computing; 2025. Available from: https://www.R-project.org/

17. Posit team. RStudio: Integrated development environment for r [Internet]. Boston, MA: Posit Software, PBC; 2025. Available from: http://www.posit.co/

18. Viechtbauer W. Conducting meta-analyses in R with the metafor package. Journal of Statistical Software. 2010;36(3):1–48.

19. Carpenter B, Gelman A, Hoffman MD, Lee D, Goodrich B, Betancourt M, et al. Stan: A probabilistic programming language. J Stat Softw [Internet]. 2017;76. Available from: https://www.ncbi.nlm.nih.gov/pubmed/36568334

20. Gabry J, Češnovar R, Johnson A, Bronder S. Cmdstanr: R interface to ‘CmdStan’ [Internet]. 2025. Available from: https://mc-stan.org/cmdstanr/

21. Kelly P, Lemmens R, Weimar C, Walsh C, Purroy F, Barber M, et al. Long-term colchicine for the prevention of vascular recurrent events in non-cardioembolic stroke (CONVINCE): A randomised controlled trial. Lancet. 2024;404:125–33.

22. Li J, Meng X, Shi FD, Jing J, Gu HQ, Jin A, et al. Colchicine in patients with acute ischaemic stroke or transient ischaemic attack (CHANCE-3): Multicentre, double blind, randomised, placebo controlled trial. BMJ. 2024;385:e079061.

23. Lou N. Colchicine goes belly-up in a more definitive heart attack trial [Internet]. 2024. Available from: https://www.medpagetoday.com/meetingcoverage/tct/112644?xid=nl_mpt_Cardiology_update_2024-11-01&mh=5ea0ef63b494fbd59d59b80f4d7177fa&zdee=gAAAAABm4utWmSHJnY-b0PoghpwIdJ2Z5bp7pHCJbHd4lnSWdd-TcQH64qhAqr5vStSuTwshVLoWZmIfruyxrtdHQaON6GGWin0MsBBlzSgmQd4CbqGcFWQ%3D&utm_source=Sailthru&utm_medium=email&utm_campaign=Automated%20Specialty%20Update%20Cardiology%20BiWeekly%20FRIDAY%202024-11-01&utm_term=NL_Spec_Cardiology_Update_Active

24. Wasserstein RL, Lazar NA. The ASA’s statement on p-values: Context, process, and purpose. The American Statistician. 2016;70:2:129–33.

25. Greenland S, Senn SJ, Rothman KJ, Carlin JB, Poole C, Goodman SN, et al. Statistical tests, p values, confidence intervals, and power: A guide to misinterpretations. European journal of epidemiology. 2016;31(4):337–50.

26. Wasserstein RL, Schirm AL, Lazar NA. Moving to a world beyond “p < 0.05.” The American Statistician. 2019;73:1–19.

27. Gelman A, Stern HS. The difference between significant and not significant is not itself statistically significant. The American Statistician. 2006;60(4):328–31.

28. Smaldino PE, McElreath R. The natural selection of bad science. R Soc Open Sci [Internet]. 2016;3(9):160384. Available from: https://www.ncbi.nlm.nih.gov/pubmed/27703703

